# Corrective Re-analysis of the Alirocumab ODYSSEY Outcomes Trial Suggests the Clinical Importance of Lipoprotein(a) Remain Substantially Underestimated

**DOI:** 10.1101/2025.10.29.25338763

**Authors:** Hu Hong

## Abstract

Although Lp(a) is an established risk factor for ASCVD, our analysis indicates its importance remains substantially underestimated. Reanalyzing cardiovascular outcomes trial (CVOT) data for the PCSK9 antibody alirocumab stratified by Lp(a) quartiles, we find that approximately 70% of the observed benefit is attributable to absolute reductions in Lp(a), rather than to lowering of LDL-C. This result aligns with a prior post hoc analysis of the PCSK9 antibody evolocumab, which attributed 57% of the benefit to Lp(a) reduction. These findings challenge the prevailing assumption that the benefits of PCSK9 therapy are mediated primarily through LDL-C lowering.

The manuscript discusses the conditions under which this counterintuitive conclusion could hold. A detailed re-examination of prior lipid-lowering therapies across different mechanisms suggests that the widely accepted linear relationship between LDL-C reduction and clinical benefit is not well supported at levels below 80 mg/dL. Lp(a) likely exerts its effects predominantly in the later stages of atherosclerosis, particularly during plaque destabilization, rather than throughout the entire course of plaque development as LDL-C does; consequently, it may not exhibit the same cumulative effect as LDL-C and could therefore demonstrate clinical benefit within relatively short follow-up periods. In addition, collider bias in secondary prevention populations with ASCVD may lead to an underestimation of the risk attributable to Lp(a).

Based on the observed relationship, we project that late-stage, Lp(a)-targeted therapies could reduce the risk of major adverse cardiovascular events (MACE) by roughly 50∼60% in phase 3 trials, which would be unprecedented in prior CVOT trials. Our projection also suggests that setting a therapeutic goal of a 15∼20% reduction in MACE would confer benefit to roughly 40% of secondary-prevention patients with elevated Lp(a), well beyond the current eligibility range (13∼21%). Further health-economic modeling suggests these therapies would have would have favorable health-economic value, as numbers-needed-to-treat would substantially lower than PCSK9 agents.

## Background

Lipoprotein(a) [Lp(a)] is an independent risk factor for atherosclerotic cardiovascular disease (ASCVD) and aortic valve stenosis (Kronenberg F, 2022). Unlike low-density lipoprotein cholesterol (LDL-C), circulating Lp(a) concentrations are predominantly genetically determined and largely unaffected by lifestyle modification. Although present at substantially lower molar levels than LDL-C, Lp(a) particles are enriched in oxidized phospholipids (OxPL) that are highly pro-inflammatory and recognizable by innate immune cells, thereby promoting vascular inflammation, atherogenesis, and thrombosis (Tsimikas S, 2005). In addition, the apolipoprotein(a) [Apo(a)] moiety of Lp(a) shares high homology with plasminogen (McLean JW, 1987), potentially impairing fibrinolysis and further increasing thrombotic risk. Multiple Mendelian randomization studies have demonstrated a robust, likely causal association between Lp(a) concentration and ASCVD risk (Langsted, 2019; Madsen, 2020; Patel, 2021; Welsh, 2022; Berman, 2024).

The extent to which pharmacologic lowering of Lp(a) translates into reductions in major adverse cardiovascular events (MACE) remains uncertain. Several late-stage investigational therapies that directly target Apo(a) and reduce Lp(a) have achieved substantial decreases (70∼100%) in early-phase trials (Tsimikas, 2020; O’Donoghue M. L., 2022; Nicholls, 2025; Nissen S. E., 2025; Nissen S. E., 2024), but cardiovascular outcomes trial (CVOT) results have not yet been reported. Prior attempts to infer expected MACE reduction from Lp(a) lowering (Burgess, 2018; Parish, 2018; Lamina & Lp(a)-GWAS-Consortium, 2019; Madsen, 2020), have often relied on a strong assumption—by analogy to LDL-C—that short-term exposure reductions capture only a fraction of lifetime, cumulative risk. Whether this assumption holds for Lp(a) biology remain unclear.

Certain drug classes, including PCSK9 monoclonal antibodies and CETP inhibitors, exert broad lipid effects, lowering both LDL-C and Lp(a). The prevailing view has been that CVOT benefits for these agents are mediated chiefly by LDL-C lowering.

Notably, a post hoc analysis of **phase 3 FOURIER trial of PCSK9 antibody evolocumab** (O’Donoghue, 2019) reported that **Lp(a) lowering accounted for 57% of the variability in clinical benefit**. However, the observation that has received limited attention.

For another PCSK9 antibody, alirocumab, there is also a post hoc analysis concluded that only 13% of clinical benefit was attributable to Lp(a) reduction (Szarek, 2020). Upon reanalysis, we identify major methodological misuse in that work and find that the contribution of Lp(a) lowering to clinical benefit was substantially underestimated; **our estimates suggest a contribution closer to ∼70%, broadly consistent with the evolocumab analysis**. Collectively, these findings imply that the impact of Lp(a) on MACE risk has been materially underestimated in the current literature.

## Data Re-analysis for Lp(a) Lowering Attribution in CVOT Benefit of PCSK antibody

The prior post hoc analysis of the alirocumab ODYSSEY OUTCOMES trial (Szarek, 2020) reported event rates and biomarker changes stratified by baseline Lp(a) quartiles for the active and placebo arms, including cardiovascular event incidence, baseline Lp(a) and its absolute reduction, and LDL-C reduction (Figure2, Table1).

**Table 1.** Baseline Lp(a), median Lp(a) lowering and median LDL-C lowering in the alirocumab group, overall and by quartile of baseline lipoprotein(a). (Szarek, 2020)

|  | Baseline Lp(a)<br>mg/dL | Median Lp(a) Lowering<br>mg/dL | Median LDL-C Lowering<br>mg/dL |
| --- | --- | --- | --- |
| Quartile 1 | < 6.7 | 0.0 | -53.3 |
| Quartile 2 | 6.7 ~ 21.2 | -5.1 | -52.7 |
| Quartile 3 | 21.2 ~ 59.6 | -9.9 | -51.2 |
| Quartile 4 | > 59.6 | -20.1 | -47.2 |
| Overall |  | -5.0 | -51.3 |

**Figure 1.**
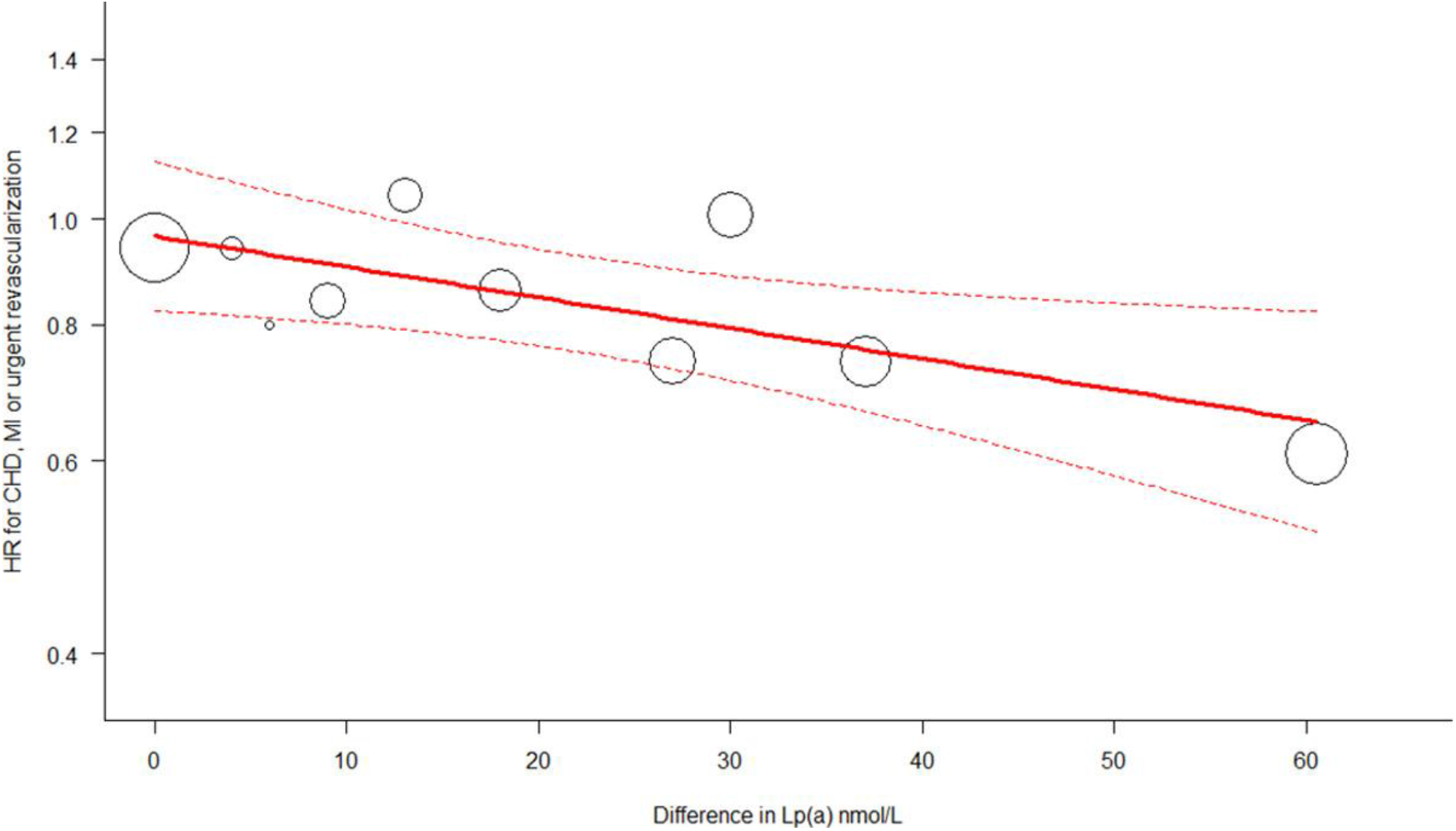
There was a significant relationship with a 15% lower risk per 25 nmol/L reduction in Lp(a) after adjusting for the change in LDL-C (model accounts for 57% of total variability of clinical benefit). (O’Donoghue M. L.-B., 2019)

**Figure 2.**
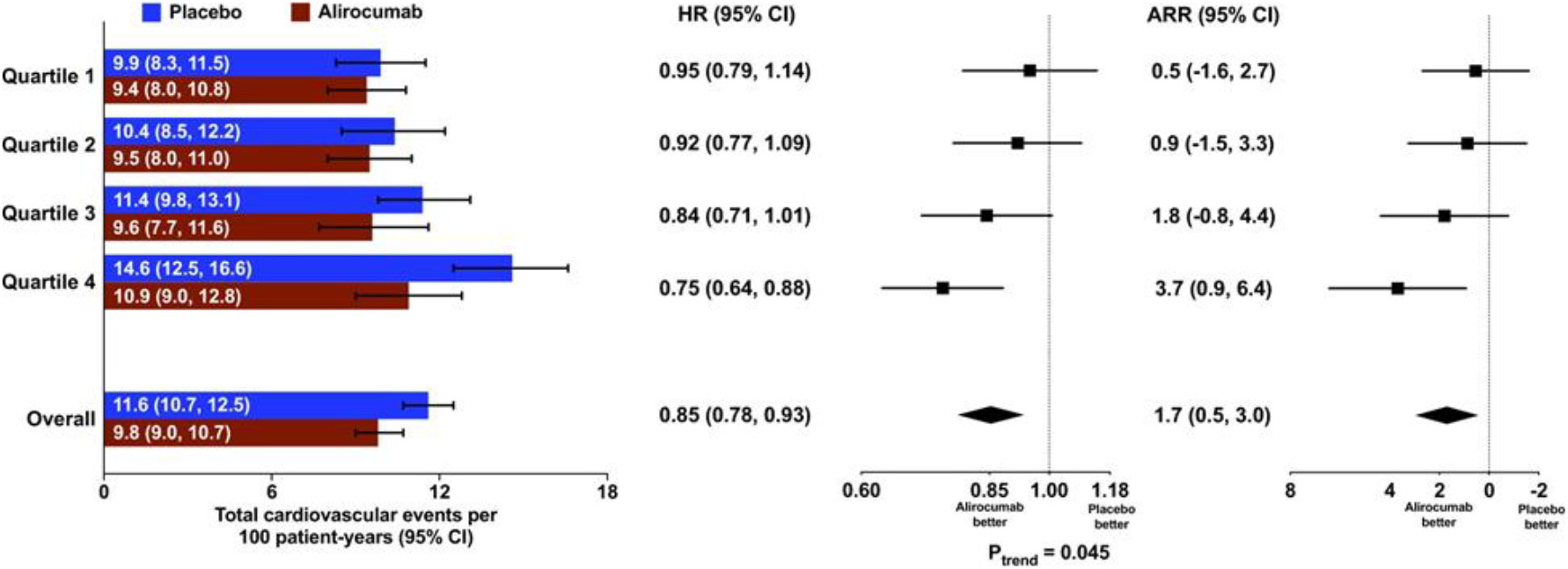
Relative and absolute treatment effects on total cardiovascular events, overall and by quartile of baseline lipoprotein(a). (Szarek, 2020)

In the lowest baseline Lp(a) quartile, the median change in Lp(a) was ∼0, whereas the median LDL-C reduction was the largest across strata. Yet the clinical benefit in this subgroup was minimal (hazard ratio [HR] ≈ 0.95; ∼5% risk reduction). Conversely, in the highest baseline Lp(a) quartile—the subgroup that also experienced the greatest absolute Lp(a) reduction—the LDL-C reduction was slightly smaller, but the observed benefit was much greater (HR ≈ 0.75; ∼25% risk reduction).

### We identified a key methodological misuse in the original analysis

The authors correlated on-treatment Lp(a) change with event counts within the treatment arm. In the unadjusted model, the correlation appeared weak; after adjusting for baseline Lp(a), a clear inverse association emerged, with little further change after additional adjustments. However, this approach primarily compares treated patients with similar baselines but different pharmacodynamic responses. What is needed for causal attribution is the relationship between absolute Lp(a) lowering and treatment effect versus placebo at a given baseline—i.e., how much of the randomized treatment benefit (active vs. placebo) is explained by Lp(a) reduction. As a result, the reported “effect of Lp(a) change” in the original work does not validly represent the proportion of cardiovascular benefit mediated by Lp(a) lowering under PCSK9 inhibition.

**We therefore re-estimated the contribution of Lp(a) lowering** using a simple mediation-style decomposition based on the quartile-level summaries, under three assumptions:

1. In the lowest baseline Lp(a) quartile, the observed HR reflects benefit attributable entirely to LDL-C lowering (i.e., negligible contribution from Lp(a) change given median ΔLp(a) ≈ 0).
2. Additivity on the log-hazard scale: The effects of LDL-C lowering and Lp(a) lowering on MACE risk are independent and additive in log space, such that for quartiles 2–4,

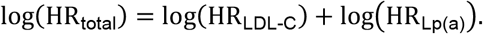
3. Approximate linearity for LDL-C: log(HR_LDL-C_) is proportional to the median LDL-C reduction across quartiles. (This assumption is not pivotal here because median LDL-C reductions are similar across quartiles.)

Operationally, we take the HR observed in quartile 1 as HR_LDL-C_. For quartiles 2–4, we compute the implied HR_Lp(a)_as

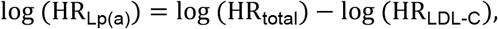

and then derive the fraction of benefit attributable to Lp(a) lowering as

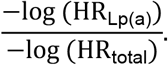

The result is shown in Table 2.

**Table 2.** Analysis result indicating the attribution of Lp(a) and LDL-C on HR, overall and by quartile of baseline lipoprotein(a).

| Quartile | $\text{HR}_{\text{Lp(a)}}$ | $\text{HR}_{\text{LDL-C}}$ | $\log(\text{HR}_{\text{Lp(a)}})$ | $\log(\text{HR}_{\text{LDL-C}})$ | Lp(a) lowering attribute | $\log(\text{HR}_{\text{Lp(a)}})$ changing per 2mg/dL Lp(a) Lowering |
| --- | --- | --- | --- | --- | --- | --- |
| Q1 | 1.000 | 0.950 | 0.000 | -0.051 | 0.0 % |  |
| Q2 | 0.968 | 0.951 | -0.033 | -0.051 | 39.2 % | -0.013 |
| Q3 | 0.882 | 0.952 | -0.125 | -0.049 | 71.7 % | -0.026 |
| Q4 | 0.785 | 0.956 | -0.242 | -0.045 | 84.2 % | -0.025 |
| Overall | 0.893 | 0.952 | -0.113 | -0.049 | 69.6 % |  |

Applying this decomposition to the quartile-stratified ODYSSEY OUTCOMES data yields a consistent pattern: the majority of treatment benefit in the higher Lp(a) strata is captured by the Lp(a)-lowering component, whereas the LDL-C–lowering component remains relatively constant across quartiles. **Aggregating across quartiles, we estimate that approximately ∼70% of the overall benefit of alirocumab is attributable to Lp(a) lowering, with the remaining ∼30% attributable to LDL-C lowering**. These results align closely with the independent post hoc findings for evolocumab highlighted a dominant role (57%) for Lp(a) reduction in explaining variability in clinical benefit.

## Why this this counterintuitive conclusion could hold

**Q1. Why would LDL-C lowering contribute little to CVOT benefit in PCSK9 trials, given LDL-C lowering is significant?**

**A. We believe that the risk slope for LDL-C flattens at very low levels**

The initial establishment of a linear relationship between LDL-C reduction and MACE risk was based on meta-analyses of statin trials (CTT, 2010), and subsequent cross-drug class meta-analyses (Silverman, 2016; Nelson Wang, 2020) further supported the consensus that this linear association holds across different therapeutic modalities. However, we believe there are multiple reasons that do not support extrapolating this linear relationship to PCSK9 antibodies.

Firstly, although trials enrolling patients with baseline LDL-C levels of 90–100 mg/dL are not uncommon, **only a very limited number of studies have achieved the LDL-C levels observed in PCSK9 trials** (30–40 mg/dL). Intensive statin therapy reduced LDL-C to approximately 60 mg/dL (PROVE-IT TIMI 22), while ezetimibe achieved 53.7 mg/dL (IMPROVE-IT). Only the CETP inhibitor Anacetrapib reached 38 mg/dL (REVEAL). This suggests that **the evidence supporting incremental clinical benefit from further reducing LDL-C from 50∼60 mg/dL to 30∼40 mg/dL remains very limited**.

Secondly, except PCSK9 trials, **clinical trials conducted in patients with relatively low baseline LDL-C have generally demonstrated lower-than-expected efficacy**. IMPROVE-IT showed only a 6% relative risk reduction, ACCELERATE showed no benefit, and REVEAL demonstrated only a 9% benefit. Looking further back, earlier intensive statin trials—including A to Z, IDEAL, and SEARCH—also failed to achieve statistical significance. However, in most of these trials, LDL-C reductions remained substantial. In summary, **at lower baseline LDL-C levels, the incremental benefit of additional LDL-C lowering seems limited**.

These analysis further reinforces our hypothesis that, in statin-treated patients whose LDL-C–mediated risk has already been substantially suppressed, non–LDL-C pathways (e.g., Lp(a)-related mechanisms) may dominate the residual benefit signal under PCSK9 inhibition.

**Q2. Many other studies** (Burgess, 2018; Parish, 2018; Lamina & Lp(a)-GWAS-Consortium, 2019; Madsen, 2020), **have estimated the magnitude of Lp(a) reduction required to achieve a given degree of risk reduction, and their estimates differ substantially from yours. What accounts for the main discrepancy? Why do you believe your estimate is the correct one?**

**A2. These studies all implicitly rely on a strong assumption: that Lp(a), like LDL-C, exerts a cumulative effect. If this assumption does not hold, then the conclusions of these studies would be fundamentally flawed**.

These analyses typically use Mendelian randomization data for Lp(a), and map it onto Mendelian or clinical trial data for LDL-C, in order to estimate how much Lp(a) reduction is equivalent to a one-unit reduction in LDL-C.

It is well established that LDL-C exhibits a cumulative effect: the risk of MACE attributable to LDL-C increases continuously with the duration of exposure to elevated LDL-C, and the benefits of LDL-C-lowering therapies likewise accrue over time with longer treatment duration. The essence of this cumulative effect, is that **LDL-C drives the entire course of atherosclerosis (i.e**., **plaque growth), a process that may span years or even decades**.

However, based on its underlying biology, we believe **Lp(a) plays a more prominent role only in the later stages of atherosclerotic disease**, particularly in plaque destabilization and rupture.

Imaging studies have shown that:

Elevated Lp(a) levels were associated with higher prevalence of presence of lipid-rich necrotic core (LRNC) and thin-or-ruptured fibrous cap (TRFC), but no association between Lp(a) and maximum vessel wall area which reflects the total plaque burden (Dam-Nolen, 2021);

In patients with an Lp(a) level of 70 mg/dL or higher, increased progression of only low-density noncalcified plaque volume are found, but no differences in baseline plaque volume nor in overall plaque progression were observed between the patients with high and low Lp(a) levels (Kaiser, 2022);

Compared with an Lp(a) level of <30 mg/dL, an Lp(a) level of ≥30 mg/dL was significantly associated with new-onset vulnerable plaque but not with stable plaque (Duan, 2023).

These findings strongly suggests that **Lp(a) likely plays a limited role in driving disease progression during the earlier stages of atherosclerotic disease, so there may not be the same decades-long cumulative effect characteristic of LDL-C**. Therefore, the strong assumption adopted in prior studies may therefore be invalid.

In contrast, during the few years preceding a MACE event, Lp(a) may exert a more pronounced effect by amplifying inflammation and promoting plaque destabilization and rupture. Under this assumption, **within the typical follow-up period of clinical trials, reducing Lp(a) could meaningfully slow plaque destabilization and thereby substantially reduce MACE incidence during the observation window**.

A further implication is that, beyond a certain treatment duration, the magnitude of benefit may plateau rather than continuing to accrue over time as observed with statins. However, this does not imply that treatment should be discontinued, as cessation would likely lead to a rebound in Lp(a) levels and a corresponding rapid increase in risk.

## Projected MACE Benefit of Lp(a)-Targeted Therapies

If our inference is correct, reductions in Lp(a) exert a decisive influence on residual risk in patients already receiving intensive statin therapy. As noted above, the empirical relationship between Lp(a) lowering and changes in log (HR_Lp(a)_)exhibits similar slopes in quartiles 3 and 4, with a smaller slope in quartile 2. This pattern implies diminishing marginal benefit when baseline Lp(a) is low, whereas an approximately linear relationship holds when baseline Lp(a) is sufficiently high. Conservatively, we assume linearity for patients whose on-treatment Lp(a) remains ≥30 mg/dL (≈72 nmol/L).

Using a working slope of Δlog (HR_Lp(a)_) ≈ 0.025 per 2 ng/dL (≈5 nmol/L) decrement in Lp(a), the implied magnitudes of MACE risk reduction (1 − HR) are shown in Table 3.

**Table 3.** Calculated magnitudes of MACE risk reduction related to Lp(a) Lowering.

| MACE reduction | Lp(a) Lowering mg/dL | Lp(a) Lowering nmol/L |  | MACE reduction | Lp(a) Lowering mg/dL | Lp(a) Lowering nmol/L |
| --- | --- | --- | --- | --- | --- | --- |
| 15% | 13.5 | 32.5 |  | 40% | 42.6 | 102.2 |
| 20% | 18.6 | 44.6 |  | 50% | 57.8 | 138.6 |
| 25% | 24.0 | 57.5 |  | 60% | 76.4 | 183.3 |
| 30% | 29.7 | 71.3 |  | 70% | 100.3 | 240.8 |

**Given that several Lp(a)-targeted agents in late-stage development produce on the order of 150–200 nmol/L reductions in Lp(a), these estimates imply approximately 50–60% relative reductions in MACE—an effect size unprecedented in prior CVOTs**.

Pelacarsen, the ASO program of Novartis & Ionis, is now in phase 3. In consideration of the phase 3 regimen (80 mg Q4W (Cho, 2025)), the anticipated degree of Lp(a) lowering likely lies between the phase 2 results for 20 mg QW (−80%) and 60 mg Q4W (−72%) (Tsimikas, 2020). If the baseline Lp(a) remains the same as in the Phase 2 trial (224.3 nmol/L), a reasonable assumption is that the Phase 3 trial will achieve a 75% reduction. Under this assumption, the expected on-treatment Lp(a) level would be approximately 56 nmol/L. As above, we conservatively assume no incremental benefit below 72 nmol/L, so the “effective” decrease is ∼152 nmol/L, corresponding to an estimated ∼53% reduction in MACE risk.

For olpasiran, phase 3 siRNA program of Amgen/Arrowhead, the median baseline Lp(a) was 260.3 nmol/L and on-treatment levels approached zero in its phase 2 (O’Donoghue M. L., 2022). Applying the same conservative threshold (no benefit <72 nmol/L), the effective decrease is ∼188 nmol/L, corresponding to an estimated ∼60% reduction in MACE risk.

Together, these projections suggest that dedicated Lp(a)-lowering agents could deliver clinically transformative—and potentially category-defining—reductions in ASCVD events, particularly among patients with substantially elevated baseline Lp(a).

## Potential of Broadening Eligible Population

If a more modest therapeutic target of 15%–20% MACE reduction is adopted, the corresponding target population can be recalculated.

We continue to make the conservative assumption that reducing Lp(a) below 72 nmol/L (approximately 30 mg/dL) would not confer additional clinical benefit; all Lp(a)-targeted therapies currently in development are capable of reaching this threshold. Based on the foregoing calculations, the target population would therefore be defined as patients with a baseline Lp(a) level of at least 105∼117 nmol/L (approximately 44∼49 mg/dL).

Using representative distributions of Lp(a) in secondary-prevention ASCVD cohorts (Figure 3.1&3.2), these thresholds imply a substantial expansion of the treatable population. Specifically, **the proportion of secondary-prevention patients who could plausibly benefit from Lp(a)-lowering therapy would increase from the oft-cited ∼13%–21% to roughly ∼40%**, based on our calculations. This shift suggests that even conservative efficacy goals could materially broaden clinical applicability and public-health impact.

**Figure 3.1 & 3.2.**
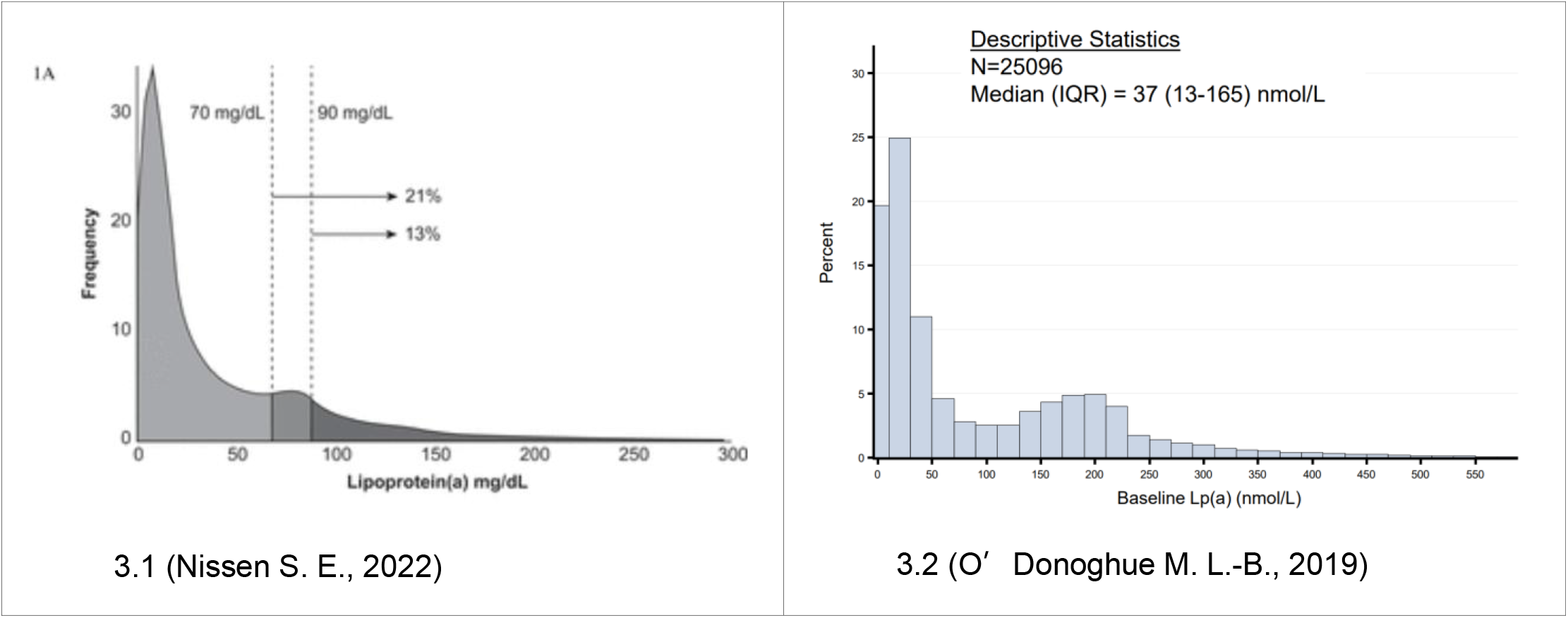
Lp(a) Distribution in secondary-prevention ASCVD cohorts

## Collider bias

**Q3**. Under your assumptions, it seems high–Lp(a) patients treated with Lp(a)-lowering therapeutics, would end up with *lower* risk than low–Lp(a) patients, which seems impossible. For example, among control group of FOURIER, **patients in the highest Lp(a) quartile (>165 nmol/L) had only ∼33% higher risk than the lowest quartile; doesn’t that cap the attainable risk reduction of lowering Lp(a) at ∼25% (0.33/1.33)** (O’Donoghue M. L.-B., 2019)?

### A. This apparent paradox arises from collider bias

In the lowest Lp(a) quartile, the randomized HR was ≈0.95, indicating only ∼5% of risk was modifiable by PCSK9 (via either LDL-C or Lp(a))—consistent with very low baseline Lp(a) and already very low on-treatment LDL-C. Inevitably, a third category of risk (neither LDL-C– nor Lp(a)–related; e.g., inflammatory, thrombotic, microvascular, or other mechanisms) accounts for the remaining ≈95% of events in that quartile. This third-risk component is independent of LDL-C and Lp(a) in the general population but jointly influences overall MACE risk.

PCSK9 CVOTs enroll secondary-prevention patients on statin therapy—i.e., individuals pre-selected for high absolute risk. When multiple independent determinants (e.g., Lp(a) and the third-risk component) influence a downstream variable (risk) that is conditioned upon by study design (selecting high-risk patients), spurious correlations (often negative) can emerge within the selected sample—the hallmark of collider bias.

**We infer that, due to this bias, Lp(a) and the third-risk component become negatively correlated in PCSK9 trial populations**: in the lowest Lp(a) quartile, the third-risk component explains ∼95% of events, whereas in the highest Lp(a) quartile its share is much less than 0.95/1.33, and the Lp(a)-related contribution is much greater than 0.33/1.33. Consequently, **the theoretical ceiling on treatment benefit from Lp(a) lowering is not constrained** by 0.33/1.33 ≈ 25%. In high–Lp(a) strata where Lp(a) contributes a larger share of total risk, substantial (>25%) risk reductions remain biologically and statistically plausible when Lp(a) is effectively lowered.

## Conclusion

Our reanalysis of PCSK9 outcomes data indicates that the clinical benefit traditionally attributed to LDL-C lowering has been materially overestimated in statin-treated, secondary-prevention populations, and that Lp(a) reduction accounts for the majority (∼70%) of observed benefit, consistent with independent findings for evolocumab (∼57%). Methodologically, reframing effect attribution on the log-hazard scale and comparing treatment vs. placebo within baseline strata resolves contradictions in prior post-hoc work and highlights a coherent, biologically plausible role for Lp(a) as a decisive driver of residual ASCVD risk. Leveraging the empirically derived slope between absolute Lp(a) lowering and log (HR_Lp(a)_), we project that late-stage, Lp(a)-targeted agents can deliver ∼50–60% reductions in MACE among patients who remain ≥72 nmol/L on treatment— magnitudes not seen in previous CVOTs. Even conservative targets (15–20% MACE reduction) would broaden eligibility to ∼40% of secondary-prevention patients with elevated Lp(a), substantially expanding the potential population beyond current expectations. Taken together, these results argue for re-prioritizing Lp(a) in risk assessment and trial design, and they support the clinical and health-economic rationale for dedicated Lp(a)–lowering therapies.

## Data Availability

All data produced in the present work are contained in the manuscript

